# Differences in early life cognitive function explain the association between low education and early dementia risk

**DOI:** 10.1101/2024.06.15.24308968

**Authors:** Bernt Bratsberg, Anders M Fjell, Ole J. Rogeberg, Vegard F. Skirbekk, Kristine B Walhovd

## Abstract

Major initiatives are currently attempting to prevent dementia by targeting modifiable risk factors. Low education is frequently pointed to as a potential key factor, due to its robust relationship with dementia risk. Impact of education is notoriously difficult to assess, however, because of associations with multiple other risk and protective factors, and large population-representative samples are required to tease the relationships apart. Here, we studied 207,814 Norwegian men born between 1950 and 1959 who underwent compulsory cognitive testing during military conscription as young adults, to systematically test associations of education, cognition, and other potentially important factors. While low education was associated with increased risk for dementia diagnosis (Hazard ratio [HR] = 1.37, CI: 1.17-1.60), this association was fully explained by earlier cognitive test scores (HR = 1.08, CI: .91-1.28). In contrast, low cognitive score was associated with double risk of later dementia diagnosis, even when taking education into account (HR = 2.00, CI: 1.65-2.42). This relationship survived controlling for early-life socioeconomic status and was replicated within pairs of brothers. The latter finding suggests that genetic and environmental factors shared within families, such as common genetics, parental education, childhood socioeconomic status, or other shared experiences, cannot account for the association. Rather, independent, non-familial factors are more important. In contrast, within-family factors accounted for the relationship between low education and diagnosis risk. In conclusion, implementing measures to increase cognitive function in childhood and adolescence appears to be a more promising strategy for reducing dementia burden.

## Introduction

Lower education is associated with increased dementia risk (1). Consistent with a view of education as a causal protective factor, dementia incidence has declined as educational attainment has increased (2), and the association does not appear to reflect differences in life-style factors, general somatic or psychiatric health, or known genetic risks (3-5). According to the Lancet commission for dementia prevention, 7% of dementia cases could be avoided by targeting low education, placing it second on the list of modifiable risk factors (6). Research using compulsory schooling reforms as natural experiments has suggested causal effects of education on later-life cognitive performance (7), but it is unclear whether this translates to lower dementia risk (8).

A popular hypothesis holds that education reduces cognitive decline and dementia by increasing cognitive reserve, which allows highly educated persons to sustain more brain pathology without cognitive decline (9, 10). Consistent with this view, education is associated with reduced dementia risk, but does not protect against development of neuropathology (11). An alternative hypothesis instead focuses on the relationship between education and cognitive function. If low education on average reflects lower earlier life functioning, baseline cognition will be closer to the functional threshold of a dementia diagnosis. As a result, age-normative cognitive decline will lead to dementia diagnosis at earlier ages for those with low education, without necessarily being associated with higher rates of decline. Consistent with this view, low cognitive scores for males tested at military conscription were associated with higher dementia risk decades later (12-14), and the association was only partly accounted for by family factors (14). A smaller study found even stronger relationships for women (15). Education could not explain the increased risk associated with low cognitive function in these studies, but adjustment for education has also been reported to fully account for the relationship between cognitive function and later dementia (16).

Whether the association between education and dementia exists independently of earlier cognitive function is an important question, as it speaks directly to the potential causal impact of education. Here we tested whether earlier cognitive function could account for increased dementia risk in people with low education. Using administrative register data with 77.4% population coverage, we studied 207,814 men born in Norway between 1950 and 1959 with cognitive test scores from compulsory military conscription and dementia diagnoses available from the health care system, in the age range 60 to 69 years (see Table S1). This captures relatively early cases of dementia, an important and under-studied group experiencing a substantial reduction in years of healthy living. Although the genetic component is important, familial autosomal dominantly inherited Alzheimer’s disease (AD) accounts for a small proportion of all early-onset cases (<0.1%) and other genetic and environmental factors need to be considered (5, 17).

## Results

Participants were categorized in five educational (compulsory, low secondary, high school, college, postgraduate, Figure S1) and seven cognitive levels (Figure S2), with the middle category as reference. Percentage of the sample with dementia diagnosis as a function of age is shown in Figure S3. Cox Proportional Hazards Model showed that low education was associated with higher risk of dementia diagnosis (Hazard Ratio [HR] = 1.37, 95% CI: 1.17-1.60, p < .001), see Figure 1. No other educational levels were associated with different risks in the sample with available cognitive scores. Results for the full population of 268,614 is included for comparison, indicating even stronger relationships. Adding cognitive scores to the model removed the association (HR = 1.08, CI: .91-1.28, p = .40), while the two lowest cognitive categories showed increased risk even when education was controlled for (lowest: HR = 2.00, CI: 1.65-2.42, p < .001; 2^nd^ lowest: HR = 1.37, CI: 1.15-1.65, p = .001). The relationships survived controlling for early-life socioeconomic status as indexed by parental education and income.

**Figure 1.**
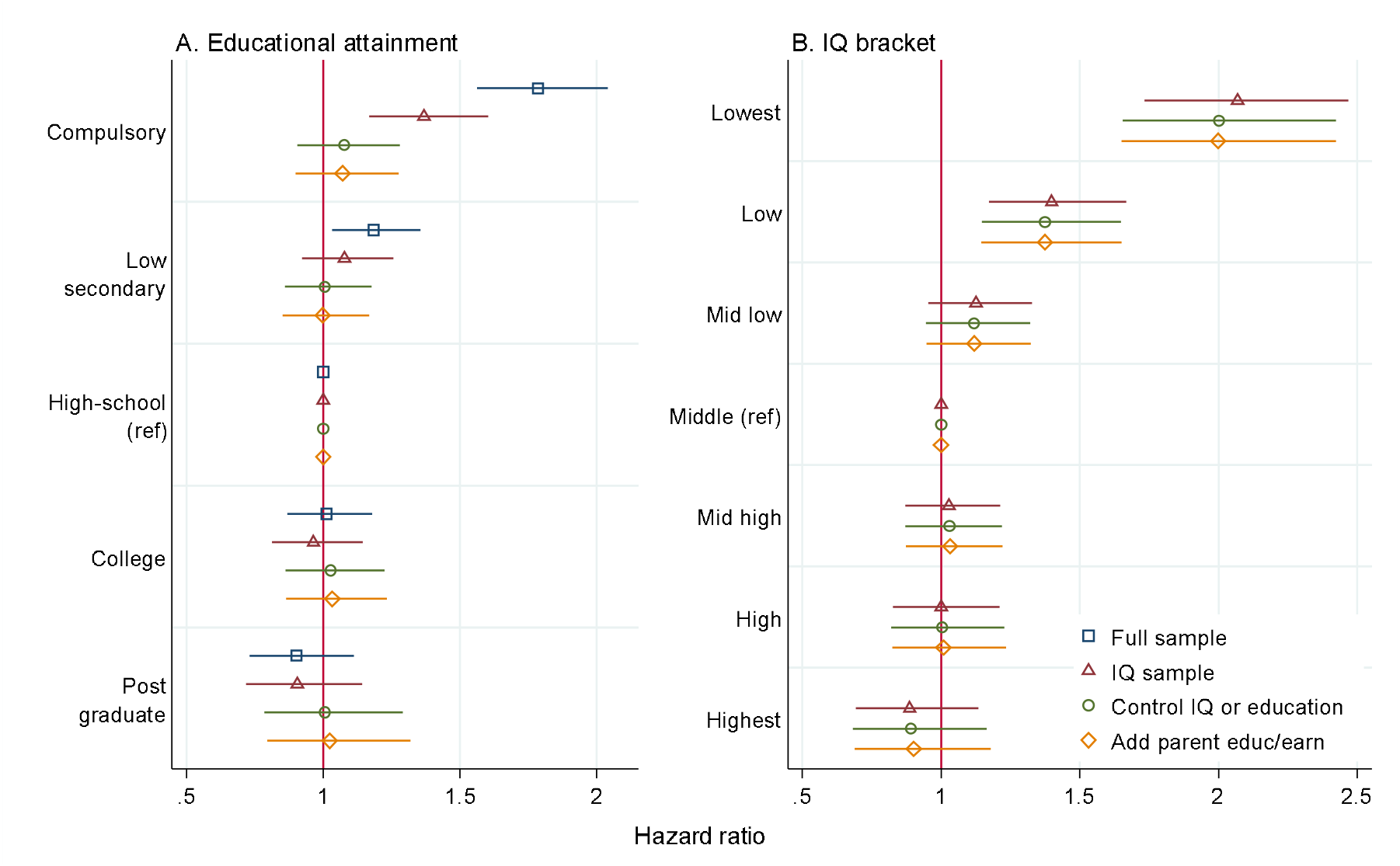
Risk of dementia diagnosis, education, and earlier cognitive function Each horizontal bar represents results controlling for different covariates. Panel A. Hazard ratios [HR] for dementia diagnosis with level of education, high school as reference category. Blue squares: baseline model, complete sample; maroon triangles: baseline model, sample with IQ score; green circles: controlling for IQ; orange diamonds: additionally controlling for parental education/earnings. Panel B. HR for diagnosis with level of cognition, medium score as reference: Maroon triangles: baseline model; green circles: controlling for education; orange diamonds: additionally controlling for parental education/earnings. Error bars denote 95% CI.

Linear probability models yielded similar results (Figure S4). There was no attenuation of effects when using within-family variation in cognitive test score to predict risk, with the point estimate for the lowest score category relative to the middle increasing from 0.70 percentage points (95% CI: 0.55 to 0.86) in the baseline model to 0.90 (CI: 0.50 to 1.30) when adding family fixed effects. For education, however, the point estimate for the lowest educational category relative to HS declined from 0.22 percentage points (CI: 0.11 to 0.33) in the baseline model to 0.05 (CI: -0.23 to 0.33), suggesting that within-family factors can account for the relationship.

Life-events were mapped as a function of cognitive scores (Figure 2). The lowest cognitive category was associated with a steep reduction in employment, from ∼90% at age 30 years to ∼50% at 60 years. Disability increased from below 20% to ∼50%, and they had fewer children, were less often married, and had a sharper increase in mortality. The 2^nd^ lowest category showed the same tendencies to a lesser extent.

**Figure 2.**
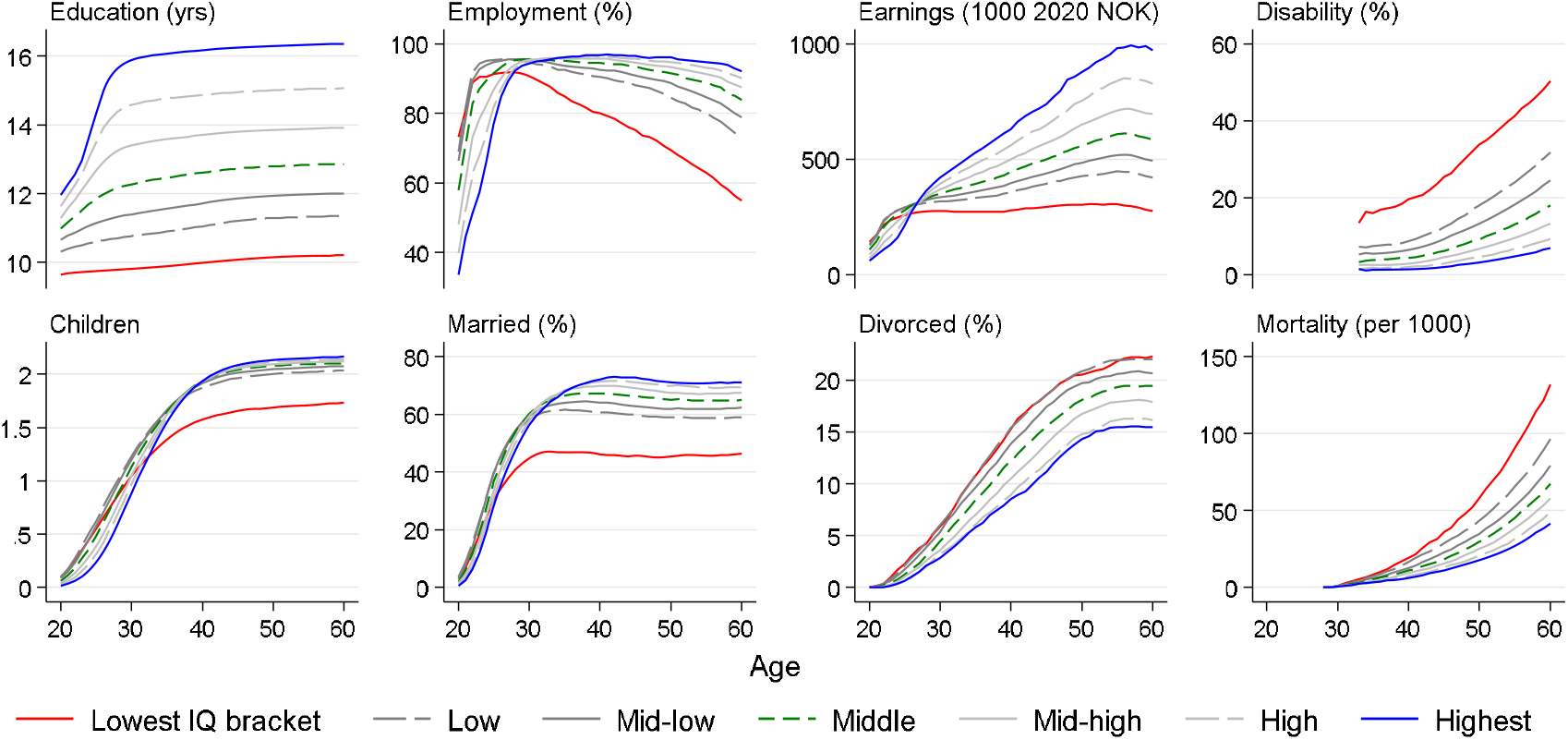
Life-events as a function of cognitive scores Except for mortality, all statistics are conditional on survival until depicted age. Disability data are available from 1992. Mortality statistics are conditional on survival until age 28.

## Discussion

Increased dementia risk associated with low education could fully be explained by earlier cognitive function. In contrast, low cognitive function was a risk factor, even when correcting for education and other factors. We show that education has no association with dementia diagnosis in this sample independently of earlier life cognition, and that suggestions of low education being a modifiable risk factor may be overly optimistic (6). The results underscore the importance of taking factors associated with early cognitive function into account for later-life diagnoses (18), which may include education prior to the age at cognitive testing (19).

Within-family variation in cognitive ability, reflecting non-shared genes or non-shared environment, showed the same association to early dementia diagnoses as seen in the full data. This means that factors shared within families, such as sibling-shared genes, childhood socioeconomic status, parental education, or other shared family experiences, cannot account for the findings. In contrast, the increased risk associated with low education did not survive controlling for within-family effects, demonstrating that shared family genetic or non-genetic factors can explain the association.

Although the mechanisms are unknown, the association may emerge because people with low cognitive scores are closer to a limit of function. As dementia diagnosis reflects loss of daily life function, diagnosis could be caused by a lower cognitive starting level, not more decline. This interpretation is in accordance with findings that education is associated with level of cognitive function but not different decline (20). The lowest cognitive category, equal to IQ < 79, had double risk of dementia diagnosis. Here, age-normative loss of function can easily be detrimental for daily life function. This is mirrored in the employment rate for this group, declining from ∼90% in young adulthood to ∼50% at age 60 years, compared to ∼75% for the 2^nd^ lowest category at this age. Accordingly, while low function was a risk, above mean score did not yield additional protection.

An alternative explanation is that people with various levels of intelligence engage in different activities and get exposed to events and environments that in turn influence dementia risk via various pathways, such as occupation, income, health, and family. Figure 2 shows that low cognitive level is indeed associated with trajectories of important life events. However, controlling for several such factors do not remove the risk (12, 21). A third theory is that cognitive level is an indicator of ‘system integrity’, reflecting how well the system is “put together”(22). Lower abilities may signify a vulnerable system, in line with the increased mortality rates. This may partly reflect earlier and even prenatal events (23), affecting cognitive function and risk for later disease.

It is important to acknowledge that associations with broad variables such as education will vary considerably across time and societies (24). Other limitations include that only men were studied and results may be different for women, a high share of all cases of dementia may never be diagnosed, which may affect the associations, and that our sample was relatively young and it is possible that other associations could be observed in older samples with higher dementia prevalence. Relatedly, data did not allow distinguishing between different types of dementia, which have different prevalence at different ages. Nevertheless, the results align with recent neuroimaging findings that brain atrophy is not related to educational level (25) but to cognitive function independently of education (26). Relationships between cognitive function and brain structure are established early (27), and are unlikely to be heavily influenced by education. Furthermore, genetic evidence has converged on cognitive function having an independent effect on AD risk, while the association with education is driven by cognition (28-30). Hence, the present results are in accordance with recent neuroscientific and genetic evidence.

## Materials and Methods

### Data

Data come from Norwegian administrative registers covering cognitive test scores, educational attainment, demographics and health care, with pseudonymous individual level identifier allowing for cross-register linkages.

#### Cognitive test scores

are from military conscription tests available in digitized form starting with the 1950 birth cohort. The stanine test score aggregate results across three speeded tests of arithmetic (30 items), word similarities (54 items), and “Raven-like” figures (36 items). The test items and scoring norms were unchanged across the study period. Coverage was high and increasing through the study cohorts, 73% for the 1950 to 86% for the 1959 cohort.

#### Educational data

come from the national education database, which contains the highest attained education drawing on administrative school records since 1974. Educational attainment brackets are given by the first digit in an education’s NUS2000 code (Norwegian Standard Classification of Education 2000).

#### Demographics

come from the population register, containing birth year and parental identifiers that allow for the identification of siblings.

#### Health care

data come from three sources. Health care in Norway is publicly financed, and the KUHR database contains data on state reimbursement starting in 2006, including diagnostic codes from all consultations with a primary physician. Patients referred to specialist care will also be observed with diagnoses in NPR (Norwegian Patient Registry) from 2008. Finally, we use diagnostic codes from the cause-of-death registry, available for the full study period.

To account for family background and family structure, we restrict the analyses to native-born individuals with two native-born parents. We further restrict our analyses to individuals with a valid education record at age 30 and, to ensure inclusion in all three health registries, present in Norway the year of the 58^th^ birthday, yielding an overall sample of 268,614, of whom 207,814 (77.4%) have a valid cognitive ability score.

### Statistical methods

Cox proportional hazard models were used for the main analyses. Observations were left censored as diagnoses prior to 2006 were not available in data, and right censored at death or if undiagnosed with dementia in the final outcome data year available (2019). Analyses were done in Stata version 17.0. Linear probability models were used to implement family fixed effect designs, using the reghdfe command in Stata.

## Data Availability

The data are parts of Norwegian national registries, and are available upon requests to the respective data owners.

## Acknowledgement

This research was supported by National Institutes of Health grant no. R01AG069109-01 (to B.B. and V.S.), the Norwegian Research Council (to K.B.W.) and UiO:Life Science (to A.M.F.).

## Supplemental Information

**Table S1.**
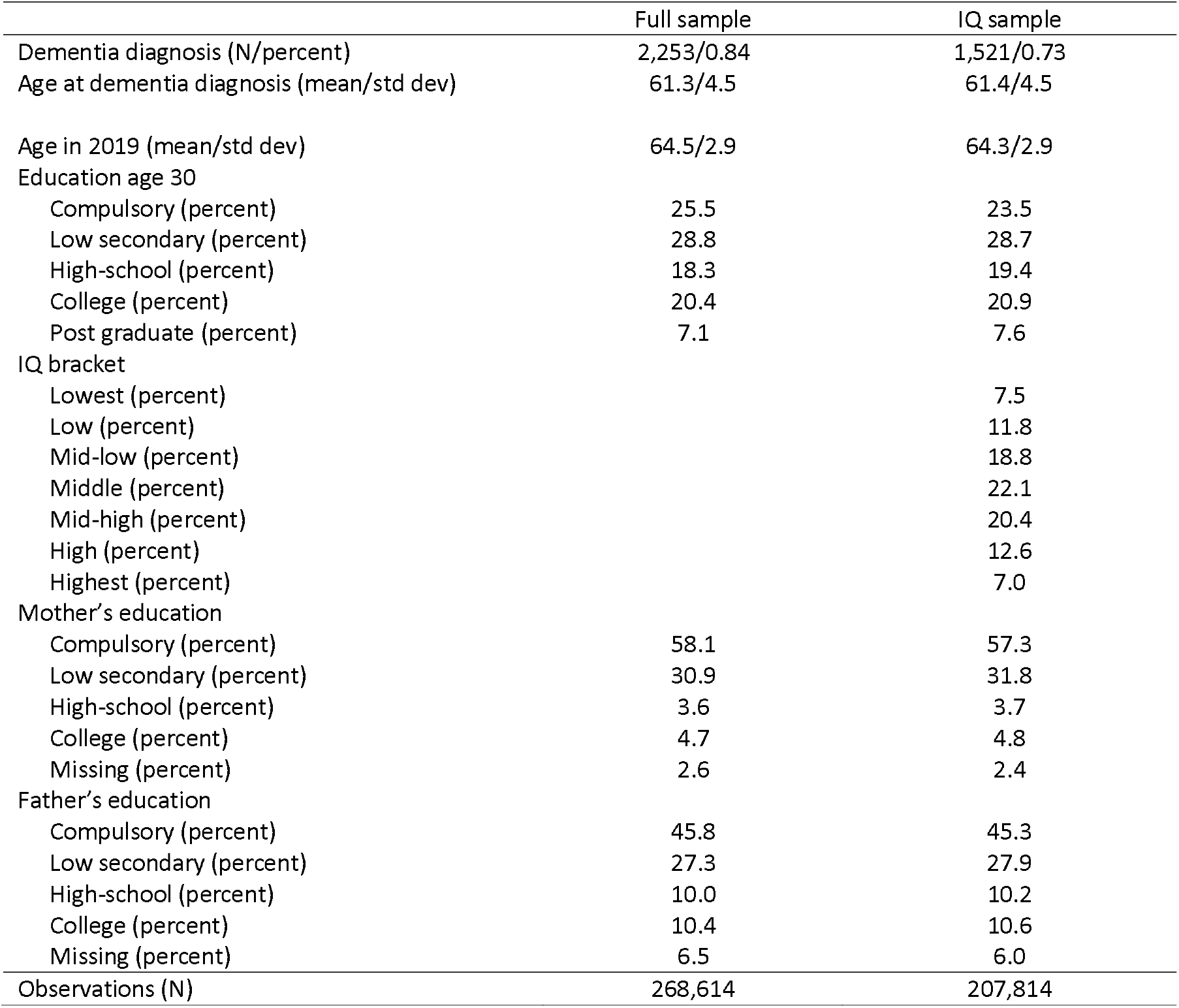
Descriptive statistics for the full sample and for the sample with cognitive scores.

**Figure S1.**
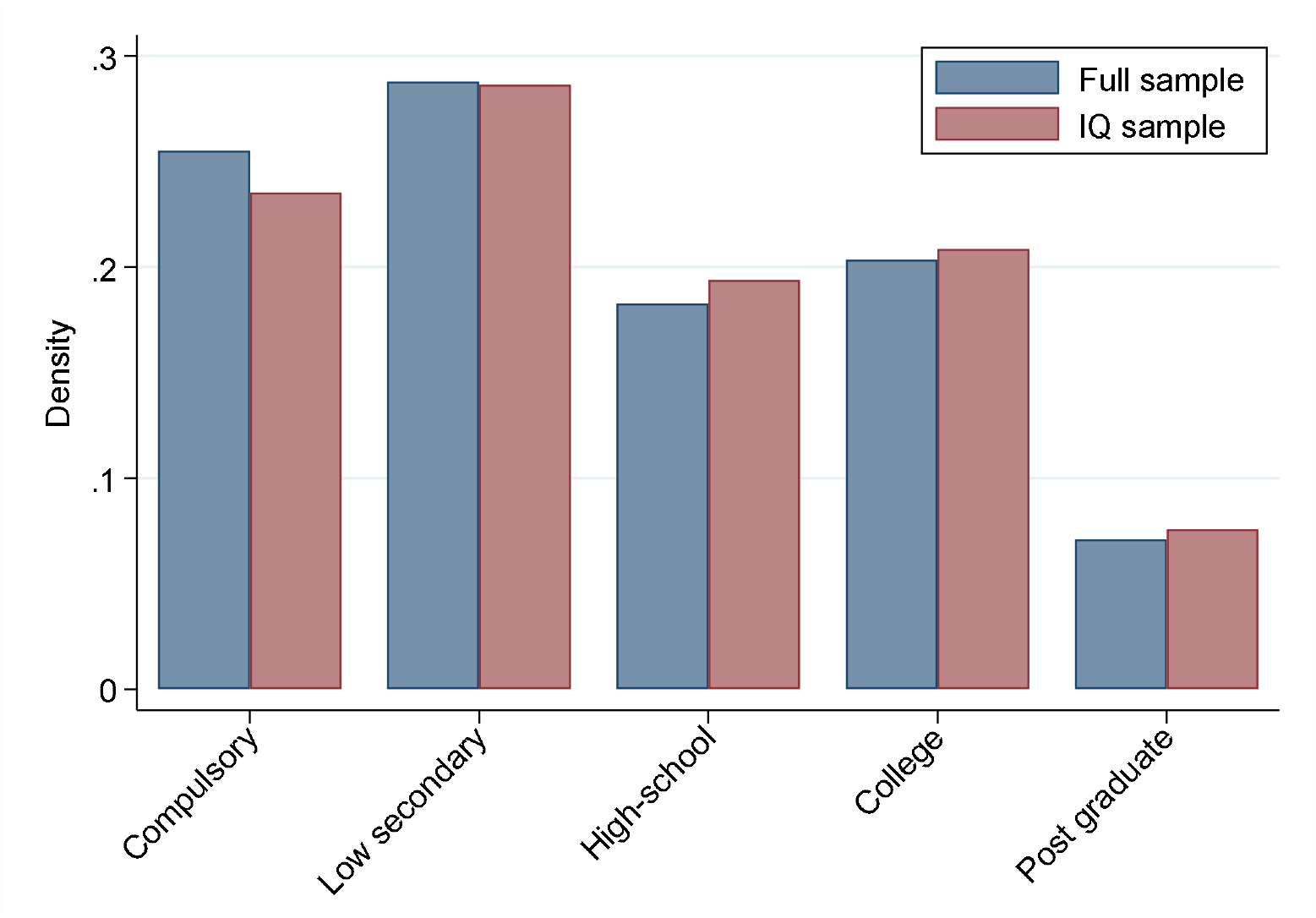
Distribution of the sample across the different educational categories. Blue bars represent the full sample (n= 268,614), maroon bars represent the sample with cognitive scores (n=207,814).

**Figure S2.**
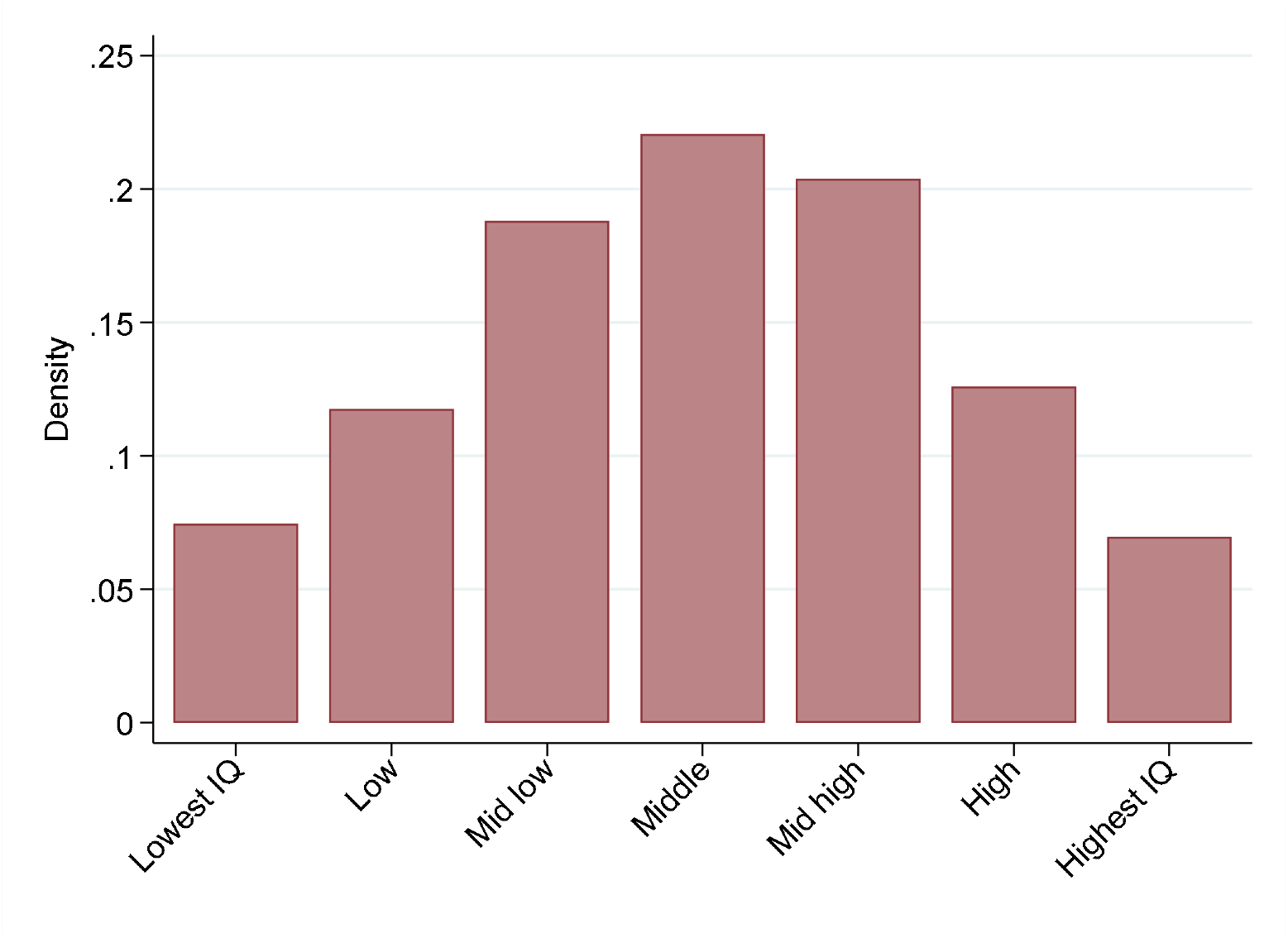
Distribution of participants across categories of cognitive scores (n= 207,814).

**Figure S3.**
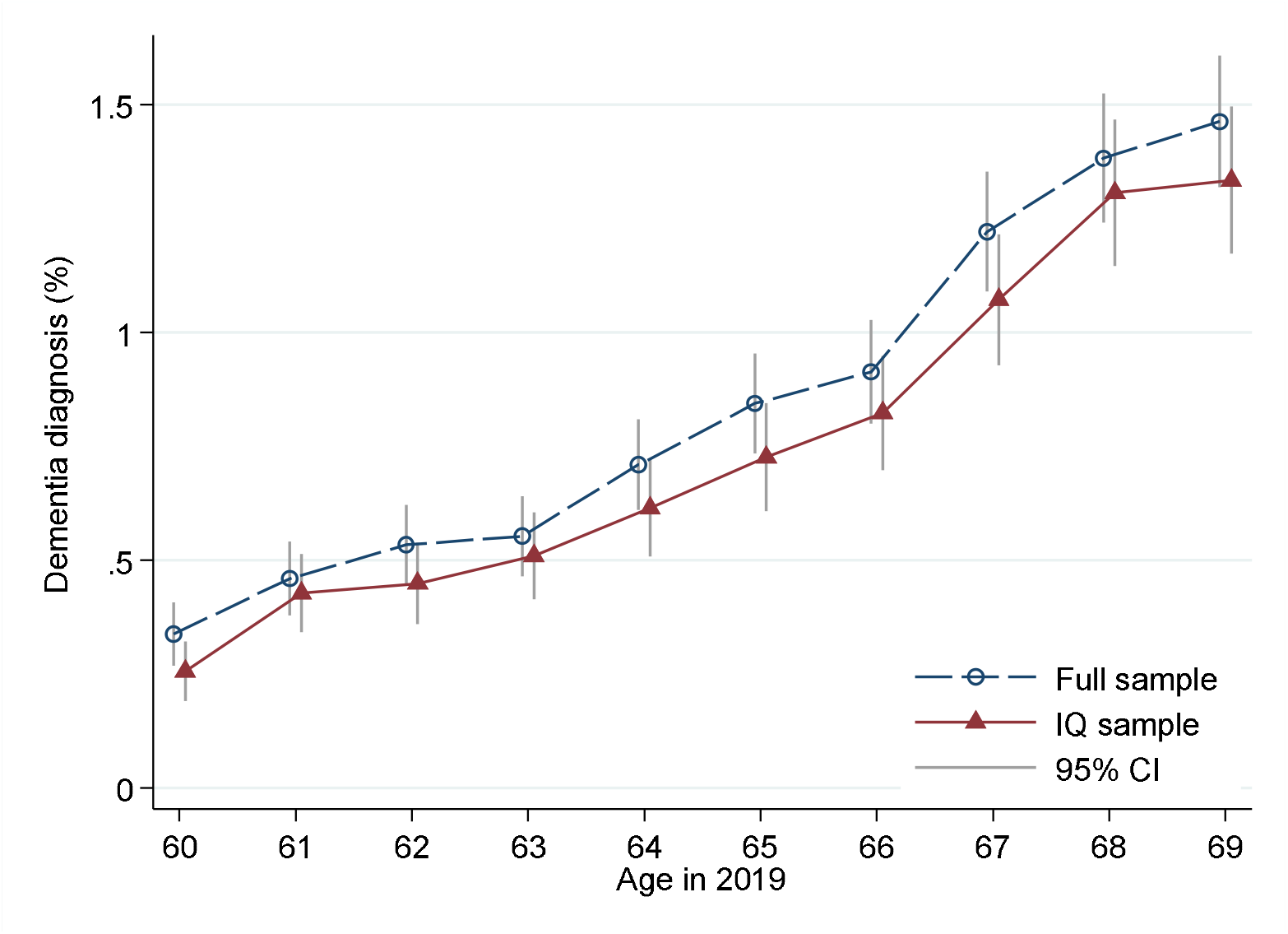
Percentage of participants with a dementia diagnosis across age. Blue circles represent the fill sample, maroon triangles represent the sample with cognitive scores. Vertical bars represent 95% CI.

**Figure S4.**
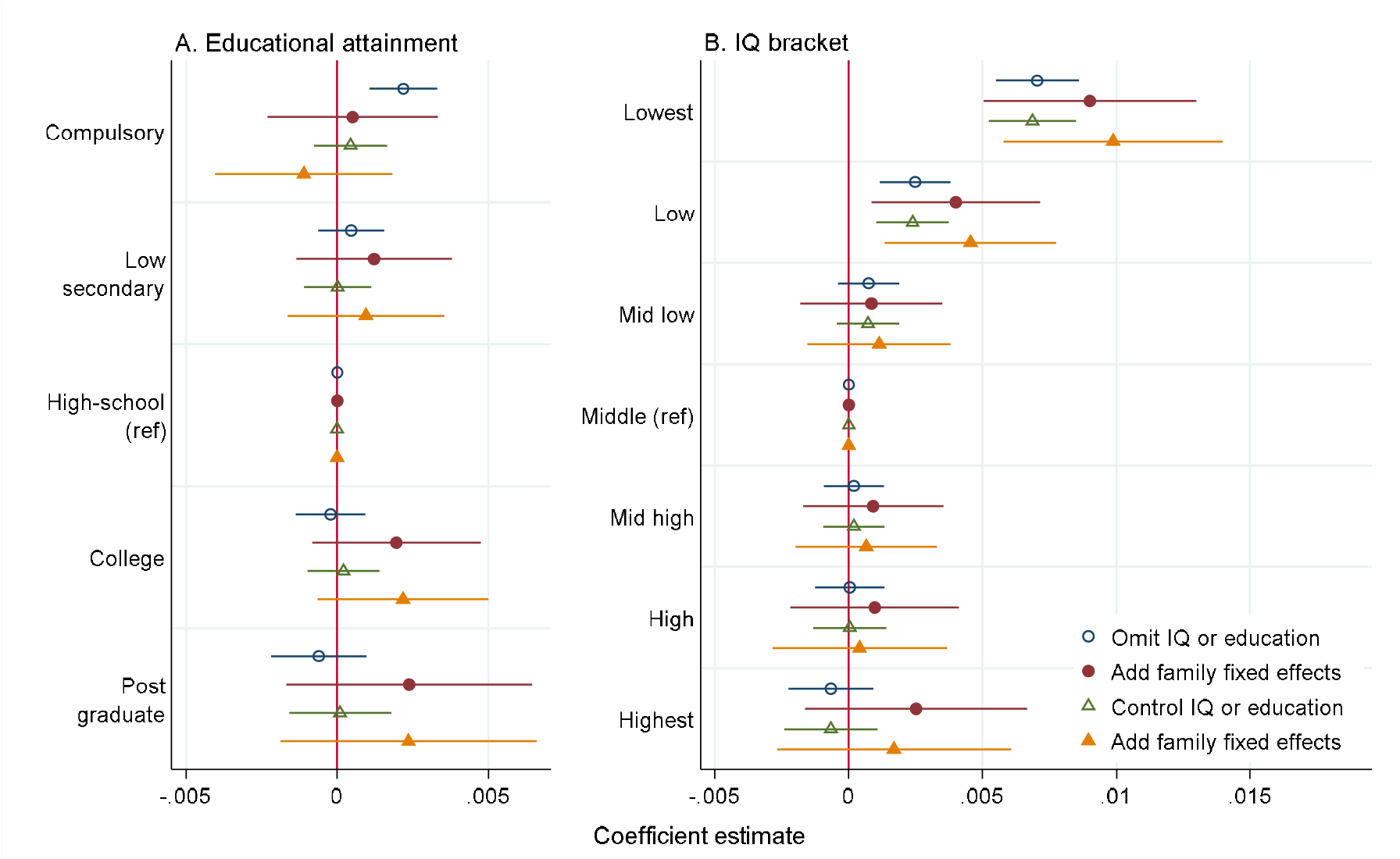
Coefficient estimates from linear probability models without and with family fixed effects Panel A. Coefficients from linear regression of dementia diagnosis on level of education, high school as reference category. Each horizontal bar represents results controlling for different covariates: Blue hollow circles: baseline model; maroon solid circles: baseline model with family fixed effects; green hollow triangles: IQ; orange solid triangles: IQ and family fixed effects. Panel B: Blue hollow circles: baseline model; maroon solid circles: baseline model with family fixed effects; green hollow triangles: education; orange solid triangles: education and family fixed effects. Error bars denote 95% CI. All models include full set of birth cohort indictor variables.

## References

1. Sharp ES & Gatz M (2011) Relationship between education and dementia: an updated systematic review. Alzheimer Dis Assoc Disord 25(4):289–304.

2. Wolters FJ, et al. (2020) Twenty-seven-year time trends in dementia incidence in Europe and the United States: The Alzheimer Cohorts Consortium. Neurology 95(5):e519–e531.

3. Ngandu T, et al. (2007) Education and dementia: what lies behind the association? Neurology 69(14):1442–1450.

4. Hendriks S, et al. (2023) Risk Factors for Young-Onset Dementia in the UK Biobank. JAMA Neurol.

5. Li R, et al. (2023) Associations of socioeconomic status and healthy lifestyle with incident early-onset and late-onset dementia: a prospective cohort study. Lancet Healthy Longev 4(12):e693–e702.

6. Livingston G, et al. (2020) Dementia prevention, intervention, and care: 2020 report of the Lancet Commission. Lancet 396(10248):413–446.

7. Schneeweis N, Skirbekk V, & Winter-Ebmer R (2014) Does education improve cognitive performance four decades after school completion? Demography 51(2):619–643.

8. Seblova D, et al. (2021) Does Prolonged Education Causally Affect Dementia Risk When Adult Socioeconomic Status Is Not Altered? A Swedish Natural Experiment in 1.3 Million Individuals. Am J Epidemiol 190(5):817–826.

9. Stern Y (2002) What is cognitive reserve? Theory and research application of the reserve concept. J Int Neuropsychol Soc 8(3):448–460.

10. Stern Y, et al. (2023) A framework for concepts of reserve and resilience in aging. Neurobiol Aging 124:100–103.

11. Members ECC, et al. (2010) Education, the brain and dementia: neuroprotection or compensation? Brain 133(Pt 8):2210–2216.

12. Nordstrom P, Nordstrom A, Eriksson M, Wahlund LO, & Gustafson Y (2013) Risk factors in late adolescence for young-onset dementia in men: a nationwide cohort study. JAMA Intern Med 173(17):1612–1618.

13. Nyberg J, et al. (2014) Cardiovascular and cognitive fitness at age 18 and risk of early-onset dementia. Brain 137(Pt 5):1514–1523.

14. Osler M, Christensen GT, Garde E, Mortensen EL, & Christensen K (2017) Cognitive ability in young adulthood and risk of dementia in a cohort of Danish men, brothers, and twins. Alzheimers Dement 13(12):1355–1363.

15. Russ TC, et al. (2017) Childhood Cognitive Ability and Incident Dementia: The 1932 Scottish Mental Survey Cohort into their 10th Decade. Epidemiology 28(3):361–364.

16. Rantalainen V, et al. (2018) Cognitive ability in young adulthood predicts risk of early-onset dementia in Finnish men. Neurology 91(2):e171–e179.

17. Whalley LJ (2001) Early-onset Alzheimer’s disease in Scotland: environmental and familial factors. Br J Psychiatry Suppl 40:s53–59.

18. Walhovd KB, Lovden M, & Fjell AM (2023) Timing of lifespan influences on brain and cognition. Trends Cogn Sci 27(10):901–915.

19. Brinch CN & Galloway TA (2012) Schooling in adolescence raises IQ scores. Proc Natl Acad Sci U S A 109(2):425–430.

20. Lovden M, Fratiglioni L, Glymour MM, Lindenberger U, & Tucker-Drob EM (2020) Education and Cognitive Functioning Across the Life Span. Psychol Sci Public Interest 21(1):6–41.

21. Reardon S (2024) Theory of sleep as a brain cleanser challenged. Science 384(6699):948.

22. Deary I (2008) Why do intelligent people live longer? Nature 456(7219):175–176.

23. Walhovd KB, et al. (2016) Neurodevelopmental origins of lifespan changes in brain and cognition. Proc Natl Acad Sci U S A 113(33):9357–9362.

24. Calandri IL, et al. (2024) Sex and Socioeconomic Disparities in Dementia Risk: A Population Attributable Fractions Analysis in Argentina. Neuroepidemiology.

25. Nyberg L, et al. (2021) Educational attainment does not influence brain aging. Proc Natl Acad Sci U S A 118(18).

26. Walhovd KB, et al. (2022) Brain aging differs with cognitive ability regardless of education. Sci Rep 12(1):13886.

27. Walhovd KB, et al. (2022) Education and Income Show Heterogeneous Relationships to Lifespan Brain and Cognitive Differences Across European and US Cohorts. Cereb Cortex 32(4):839–854.

28. Thorp JG, et al. (2022) Genetic evidence that the causal association of educational attainment with reduced risk of Alzheimer’s disease is driven by intelligence. Neurobiol Aging 119:127–135.

29. Anderson EL, et al. (2020) Education, intelligence and Alzheimer’s disease: evidence from a multivariable two-sample Mendelian randomization study. Int J Epidemiol 49(4):1163–1172.

30. Lord J, et al. (2022) Disentangling Independent and Mediated Causal Relationships Between Blood Metabolites, Cognitive Factors, and Alzheimer’s Disease. Biol Psychiatry Glob Open Sci 2(2):167–179.

